# At Home Detection of Ovarian Health Biomarker in Menstruation Blood

**DOI:** 10.64898/2025.12.18.25342545

**Authors:** Lucas Dosnon, Thomas Rduch, Salma Sherif Azer, Inge K. Herrmann

**Affiliations:** Nanoparticle Systems Engineering Laboratory, Institute of Energy and Process Engineering (IEPE), Department of Mechanical and Process Engineering (D-MAVT), ETH Zurich, Sonneggstrasse 3, 8092 Zurich, Switzerland; Nanomaterials in Health Laboratory, Department of Materials Meet Life, Swiss Federal Laboratories for Materials Science and Technology (Empa), Lerchenfeldstrasse 5, 9014 St. Gallen, Switzerland; The Ingenuity Lab, University Hospital Balgrist, Forchstrasse 340, 8008 Zurich, Switzerland; Department of Gynecology and Obstetrics (Frauenklinik), Cantonal Hospital St. Gallen (KSSG), Rorschacherstrasse 95, 9007 St. Gallen, Switzerland; Faculty of Medicine, University of Zurich, Rämistrasse 100, 8006 Zürich, Switzerland

## Abstract

Blood-based biomarkers are central to diagnostics, yet current approaches depend on invasive sampling and centralized laboratory infrastructure. At the same time, women’s reproductive health remains severely under-monitored: most clinically relevant biomarkers are rarely measured outside fertility clinics, leaving millions without accessible, continuous insight into their reproductive lifespan. Anti-Müllerian hormone (AMH), a key indicator of ovarian reserve and overall reproductive function, still requires venous blood collection and specialized analysis, creating a major barrier to early detection, routine monitoring, and population-level screening.

Here, we present a lateral flow assay (LFA) enabling direct AMH detection in unprocessed menstrual blood. The assay uses covalently conjugated 150 nm gold nanoshells to achieve sensitive colorimetric detection within the clinically relevant 0–10 ng/mL range. Results can be visually interpreted by naked-eye detection or quantified via a smartphone-based machine-learning algorithm for semi-quantitative assessment. The LFA performance correlates strongly with clinical chemistry lab-based analyses and can be seamlessly integrated into point-of-care formats, including wearable menstruation pads as well as simple dipstick tests. This technology provides a non-invasive, affordable, and robust solution for decentralized, regular monitoring of ovarian health.

## INTRODUCTION

Ovarian health is a cornerstone of reproductive health and overall well-being in women. The ovaries play a central role in menstrual cycle regulation, producing oocytes, and secreting key hormones that support numerous physiological processes.(*1–3*) Despite its paramount importance, ovarian health is often under-monitored until symptoms of dysfunction or infertility arise, resulting in high mental and socioeconomic burden. Ovarian health can be compromised by a range of conditions, including polycystic ovary syndrome (PCOS), premature ovarian insufficiency (POI), endometriosis, or ovarian cancer, all of which can impact fertility, hormone regulation, and overall well-being.(*4–9*) However, current diagnostic methods for ovarian health are often invasive, expensive, or relying on symptoms that may not manifest until the disease has progressed.(*10–12*)

Anti-Müllerian hormone (AMH) is a 140-kDa dimeric glycoprotein hormone belonging to the transforming growth factor-β (TGF-β) superfamily. (*13*, *14*) Clinically, AMH is secreted by granulosa cells of ovarian follicles and is a clinical biomarker for ovarian reserve, offering advantages over cyclically variable hormones such as Follicle-Stimulating Hormone (FSH) or estradiol due to its relative stability throughout the menstrual cycle.(*15*, *16*) AMH measurement is routinely used to assess fertility, predict ovarian response during assisted reproductive technologies (ART), and diagnose conditions such as polycystic ovary syndrome (PCOS). It is also valuable for early detection of diminished ovarian reserve, onset of menopause, and for monitoring granulosa cell tumors(*17–21*). AMH is a stable, longitudinal biomarker of ovarian health and reproductive aging, not limited to fertility assessment, enabling non-invasive, decentralized monitoring and early detection of declining ovarian function well before clinical symptoms arise. Clinically, the interpretation of AMH levels depends on multiple factors, including age, reproductive history, and clinical context. AMH levels between 1.0 and 4.0 ng/mL are generally considered indicative of a normal ovarian reserve, while values below 1.0 ng/mL may suggest diminished ovarian reserve. Values above 4.0 ng/mL are often associated with increased follicle count and may indicate ovarian conditions such as PCOS. Additionally, clinical decisions regarding fertility treatment or the risk of ovarian hyperstimulation are often based on both absolute AMH values and their interpretation within age-specific reference ranges. Therefore, a precise and accessible measurement method that can reliably quantify AMH in the 0–10 ng/mL range over time is essential for supporting fertility assessments and ovarian health evaluations across diverse female populations.(*22–24*)

While most AMH testing today relies on centralized lab-based immunoassays such as Enzyme-linked immunosorbent assay (ELISA) or chemiluminescence immunoassay (CLIA),(*25–27*) there has been attempts in extending AMH detection beyond centralized laboratory techniques. Some AMH detection approaches have been reported, but these are limited by low sensitivity, complex instrumentation as well as the requirement for pre-processed plasma or serum samples.(*28–31*) These assays exhibit limited compatibility with unprocessed whole blood, necessitating sample dilution, pre-processing steps, or the use of specialized instrumentation to achieve reliable measurements. The clinical relevance of AMH and the limitations of currently available tests highlight the urgent need for improved point-of-care AMH assays that combine high sensitivity, ease of use, and compatibility with non-invasive sample collection, critical features for expanding access to reproductive health monitoring.

Recent studies have demonstrated that menstrual blood closely correlates with venous blood for protein profiles, while also offering advantages such as reduced coagulation factor content and lower hemoglobin levels, making it particularly suitable for colorimetric paper-based tests.(*32–35*) Importantly, recent work has shown that AMH levels in menstrual blood are strongly correlated with those in venous blood,(*36*) reinforcing the potential of menstrual fluid as a clinically meaningful matrix for ovarian health monitoring. Combined with the recent integration of low-cost paper-based lateral flow assays into wearable menstruation pad sensors,(*37*) AMH monitoring in menstruation blood becomes achievable, opening new opportunities for health monitoring.

In this work, we introduce an AMH lateral flow assay employing covalently conjugated gold nanoshells and demonstrate its applicability directly in unprocessed menstrual blood, supported by a smartphone-compatible, AI-based readout system. The LFA platform can function as a dipstick or be integrated into a wearable, enabling accurate, semi-quantitative AMH detection and longitudinal monitoring without any preprocessing. Together, these features offer a powerful and accessible approach to decentralized ovarian health monitoring.

## RESULTS & DISCUSION

### Establishment of LFA for AMH detection in human serum

To enable AHM-enabled ovarian health monitoring, an LFA sensor for the detection and semi-quantification of AMH within the clinically relevant range (0–10 ng/mL) was developed, yielding a robust calibration curve that enables accurate quantification of AMH in samples of unknown concentration. First, optimization of the LFA sensor was performed to achieve detection and semi-quantification in human serum (Figure 2a). Antibody-functionalized 150 nm silica-core gold nanoshells were used as the labeling element for AMH detection (Figure 2b). During LFA development and characterization, defined volumes of AMH-spiked human serum were applied to the sample pad. The sample then migrated toward the conjugate pad, where AMH antigens interacted with the antibody-functionalized gold nanoshells pre-deposited on the pad. The resulting antigen–antibody–nanoshells complexes continued to flow toward the test line on the nitrocellulose (NC) membrane, where they were captured by immobilized antibodies, forming a sandwich complex that generated a visible signal.

**Figure 1:**
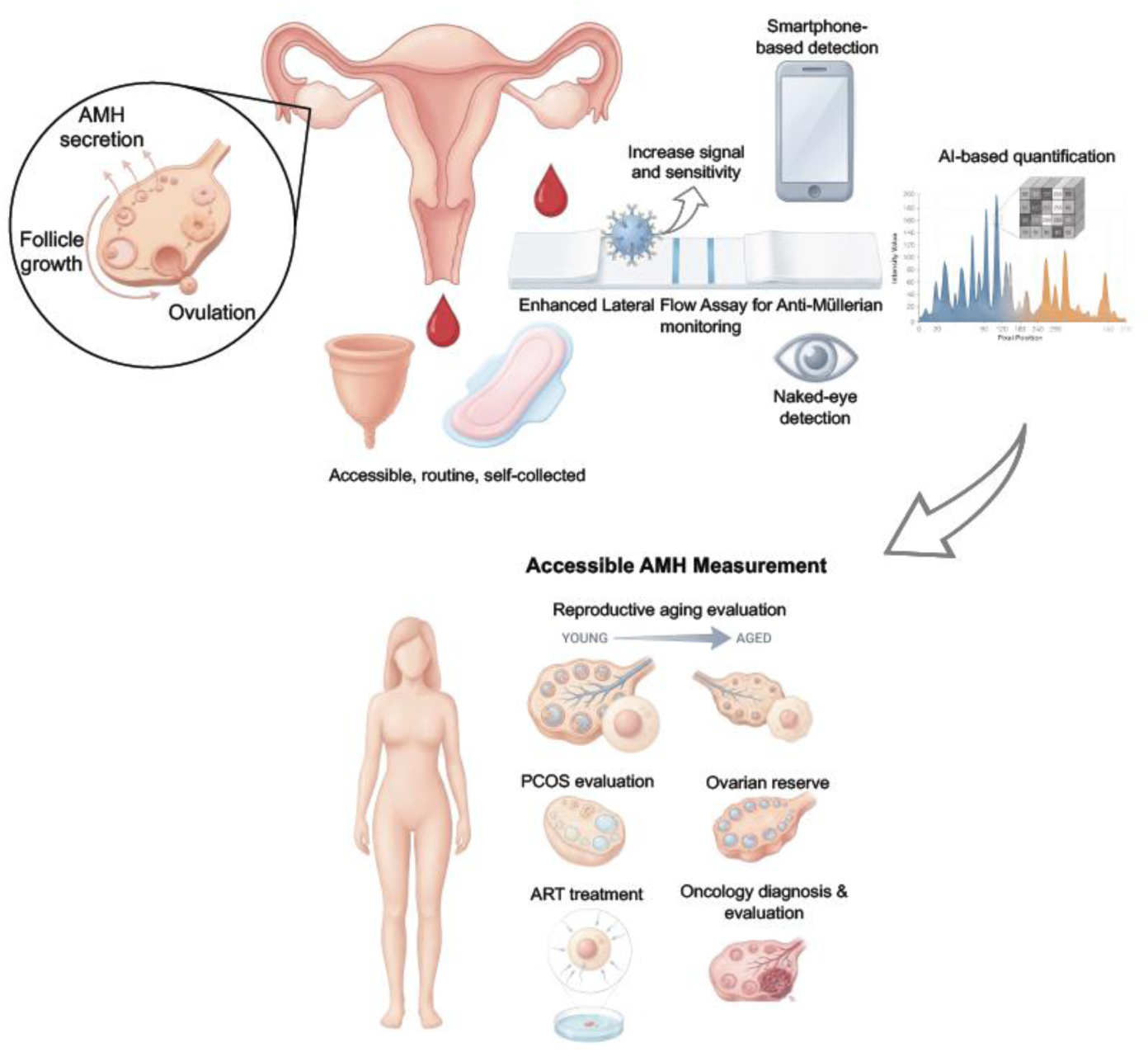
Concept of menstruation blood based anti-müllerian hormone monitoring using a point-of-care device for ovarian health evaluation. The platform includes a lateral flow assay based on 150 nm silica-gold nanoshells enabling maximized signal output and sensitivity for measurement of AMH in menstruation blood, using an AI-assisted image analysis software or naked eye.

**Figure 2:**
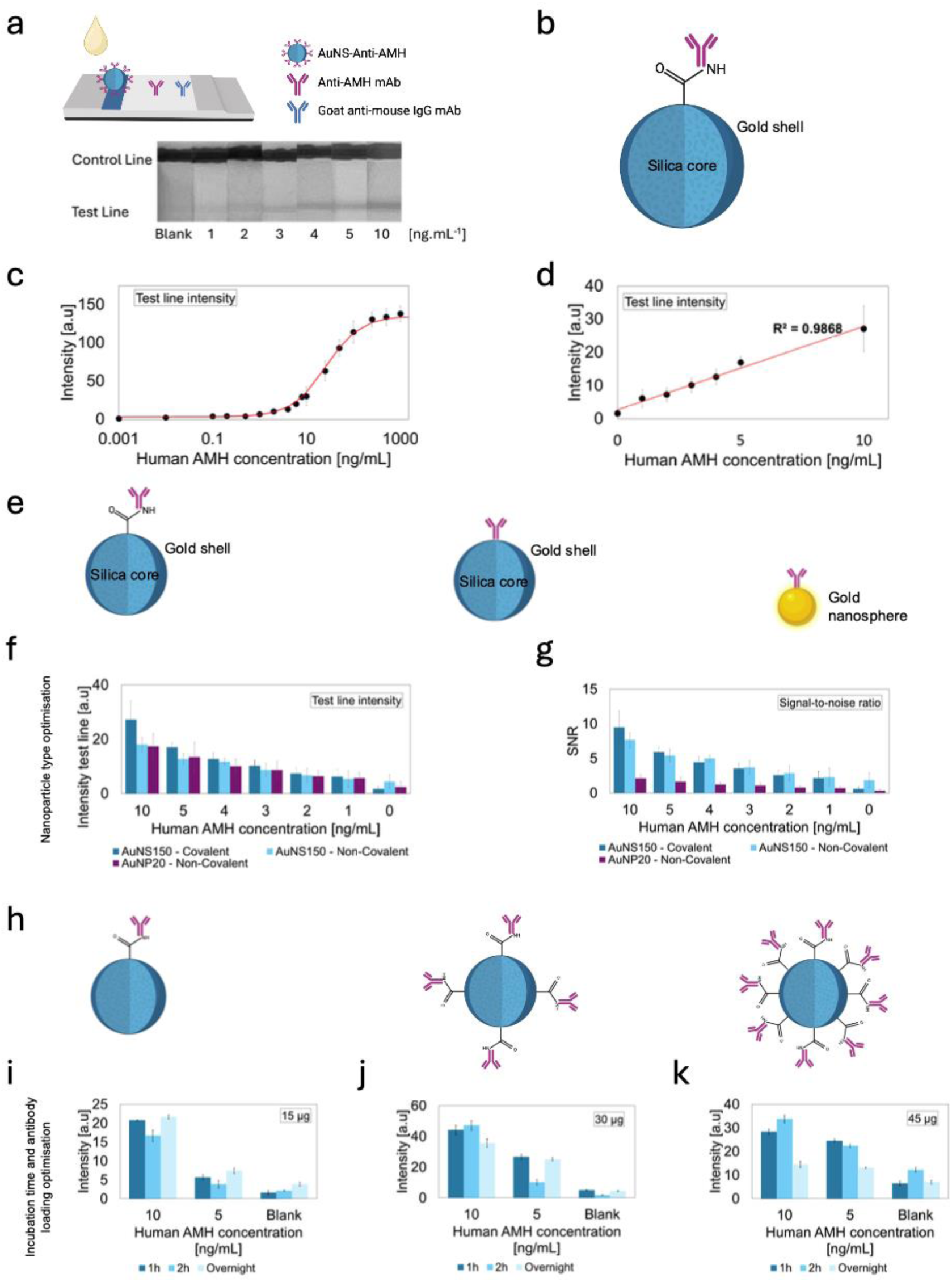
Lateral flow assay calibration curve for the detection of anti-müllerian hormone in human serum and optimization of conjugation parameters of AMH antibodies to nanoparticles. a) Design of AMH LFA assay using human AMH-antibody functionalized 150 nm silica-gold nanoshells in human serum. b) 150 nm silica gold nanoshells covalently conjugated with anti-AMH antibodies. c) Four-parameter logistic curve of signal intensity against human AMH concentration in serum. d) Linear calibration function of signal intensity against human AMH concentration in serum on the clinically relevant concentration range (0-10 ng/mL). LFA images shown in grayscale. e) Different gold nanoparticle used for optimization of AMH LFA including 20 nm gold nanoparticles and 150 nm gold nanoshells conjugated with AMH antibodies using passive electrostatic interactions and covalent conjugation. f) Test line signal intensity comparison using different nanoparticles. g) Signal-to-noise ratio comparison using different nanoparticles. h) Conjugation parameters for selection of optimal antibody concentration covalently conjugated to 150 nm gold nanoshells. i) Test line signal intensity comparison after 1h, 2h and overnight incubation of 15 µg of AMH antibodies covalently conjugated to 150 nm gold nanoshells. j) Test line signal intensity comparison after 1h, 2h and overnight incubation of 30 µg of AMH antibodies covalently conjugated to 150 nm gold nanoshells. k) Test line signal intensity comparison after 1h, 2h and overnight incubation of 45 µg of AMH antibodies covalently conjugated to 150 nm gold nanoshells.

After establishing the assay in serum, a Fusion 5 blood-separation membrane was incorporated to enable direct analysis of whole blood. This membrane efficiently retained red blood cells within its glass-fiber matrix, allowing only plasma to pass through the assay. Fusion 5 can separate up to 50 µL of blood per cm^²^ with a plasma recovery rate of 90–95%. As a result, the characteristic red coloration of whole blood was removed, enabling clear and reliable quantification of color intensity at both the test and control lines. Through analysis of the test line intensities generated at different AMH concentration, a four-parameter logistic curve (sigmoidal curve) was obtained for concentrations of AMH in human serum ranging from 0 to 1000 ng/mL (Figure 2c). The fitted curve corresponds to the following equation:

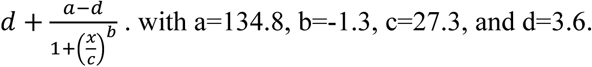

The dynamic range was obtained for concentration of AMH between 0 and 10 ng/mL, covering the clinically relevant detection window. A linear response was obtained with a correlation coefficient R^2^ of 0.98 (Figure 2d). The limit of detection (LOD) of the assay is 0.54 ng/mL. Taken together, this assay configuration demonstrates a linear response in the diagnostically important concentration range of AMH in human serum and sufficient robustness for reliable detection of AMH.

### Optimization of nanoparticle conjugates for enhanced signal output

To obtain an LFA for the detection and semi-quantification of AMH in unprocessed menstruation blood, it is crucial to ensure the best performances such as signal intensity, signal-to-noise ratio, and sensitivity, to maximize clinical utility. These performance metrics are critical for semi-quantitative detection in resource-limited and user-operated settings, especially with complex and variable matrix such as menstruation blood. Three different nanoparticle solutions were evaluated and compared to establish a performance-optimized LFA: i) 150 nm gold nanoshells (AuNS) that were covalently conjugated with anti-AMH antibodies through EDC/NHS chemistry. ii) 150 nm AuNS passively conjugated with anti-AMH antibodies through electrostatic interactions, and finally iii) 20 nm gold nanoparticles (AuNP) passively conjugated with anti-AMH antibodies through electrostatic interactions (Figure 2e).

The LFAs for the detection and quantification of AMH were prepared and tested using serum samples spiked at various concentration on the pre-determined linear range (0 to 10 ng/mL). Signal intensity and signal-to noise ratio were recorded and compared for the different nanoparticle solutions used. While 20 nm gold nanoparticles enabled sufficient signal intensity and sensitivity for the detection and semi-quantification of AMH, 150 nm gold nanoshells offered superior signal intensity, sensitivity and signal-to-noise ratio and were therefore selected. Furthermore, covalent conjugation of anti-AMH antibodies on the surface of the 150 nm gold nanoshells further enhanced the signal intensity and reduced the blank signal. Signal-to-noise ratio was also maximized (Figure 2f,g). Further testing was undertaken to fully optimize the parameters of the covalent conjugation of anti-AMH antibodies to the surface of gold nanoshells. First, different concentrations of antibodies were used; 15, 30 and 45 µg were respectively added to 1 mL of gold nanoshells for conjugation (Figure 2h). Second, different incubation times were investigated to optimize the conjugation. Antibody-nanoshells solutions were left for incubation for 1h, 2h and overnight. The signal intensity at different concentrations of AMH was recorded and compared between the different set of parameters of covalent conjugation. While the use of more antibodies seemed to enhance the signal intensity, too much of it introduced a higher signal for the blank sample coming from potential non-specific interactions between proteins. Ultimately, 30 µg of antibodies was selected as the concentration of choice for optimal assay performance. Finally, incubation times of 1h for the AuNS-antibody coupling enabled the best performance in terms of signal intensity and signal-to-noise ratio (Figure 2i,j,k). Such enhanced readout fidelity is crucial for enabling semi-quantitative detection of AMH in whole, unprocessed menstrual blood, where matrix interferences can obscure weak signals. These findings not only established the sensing foundation for our LFA but also demonstrated how particle–surface chemistry directly influences real-world usability and analytical robustness.

### Smartphone-assisted semi-quantitative analysis of anti-müllerian hormone

LFA results are prone to subjective interpretations, leading to frequent false positives or negatives. (*38*, *39*) Unprocessed whole blood can cause interferences for colorimetric readouts, caused by red blood cells leaking through separation pads, non-specific interactions, or lysed cells creating a background interference, making the interpretation and analysis of test results particularly challenging for untrained users or in the absence of assistive technologies. This is particularly critical for AMH assays, which require a high level of sensitivity and precision to ensure accurate evaluation.

To address the issues, we performed automatic image analysis using an improved machine-learning image analysis pipeline that expands previous work thanks to a more comprehensive training dataset and a redesigned peak-segmentation model to improve accuracy, robustness and signal extraction (Figure 3a)(*37*). The approach involved a smartphone-captured image of the LFA strip, and subsequent analysis with detection and segmentation models (Figure 3b). The machine-learning based software was developed using a tailored training and testing dataset, as well as a sophisticated model that allows automatic peak detection from a pixel-intensity graph obtained after automatic detection of the readout zone. The application first automatically detected the readout zone, where colorful lines (control and test) were visible. For this purpose, a pre-trained yolov8 convolutional neural network (CNN) was employed (Figure 3c), achieving a precision of 97.7%, a recall of 97.9% and an mAP50 of 0.989 (Figure 3d). Once the readout zone is accurately detected and segmented, the pixel intensity graph was extracted. It was then processed by a model built using an encoder-decoder architecture (Figure 3e), enabling peak segmentation on the intensity profile. The model could accurately detect the peaks corresponding to the intensities of the control and the test lines, with a precision of 93.7%, a recall of 94.9% and a dice score of 0.94 (Figure 3f). Once the intensity of the peak corresponding to the test line was extracted, the concentration of AMH measured by the LFA was obtained using the pre-determined calibration curve. The resulting pipeline provides a reliable and fully automated approach for interpreting AMH LFA results and is readily deployable.

**Figure 3:**
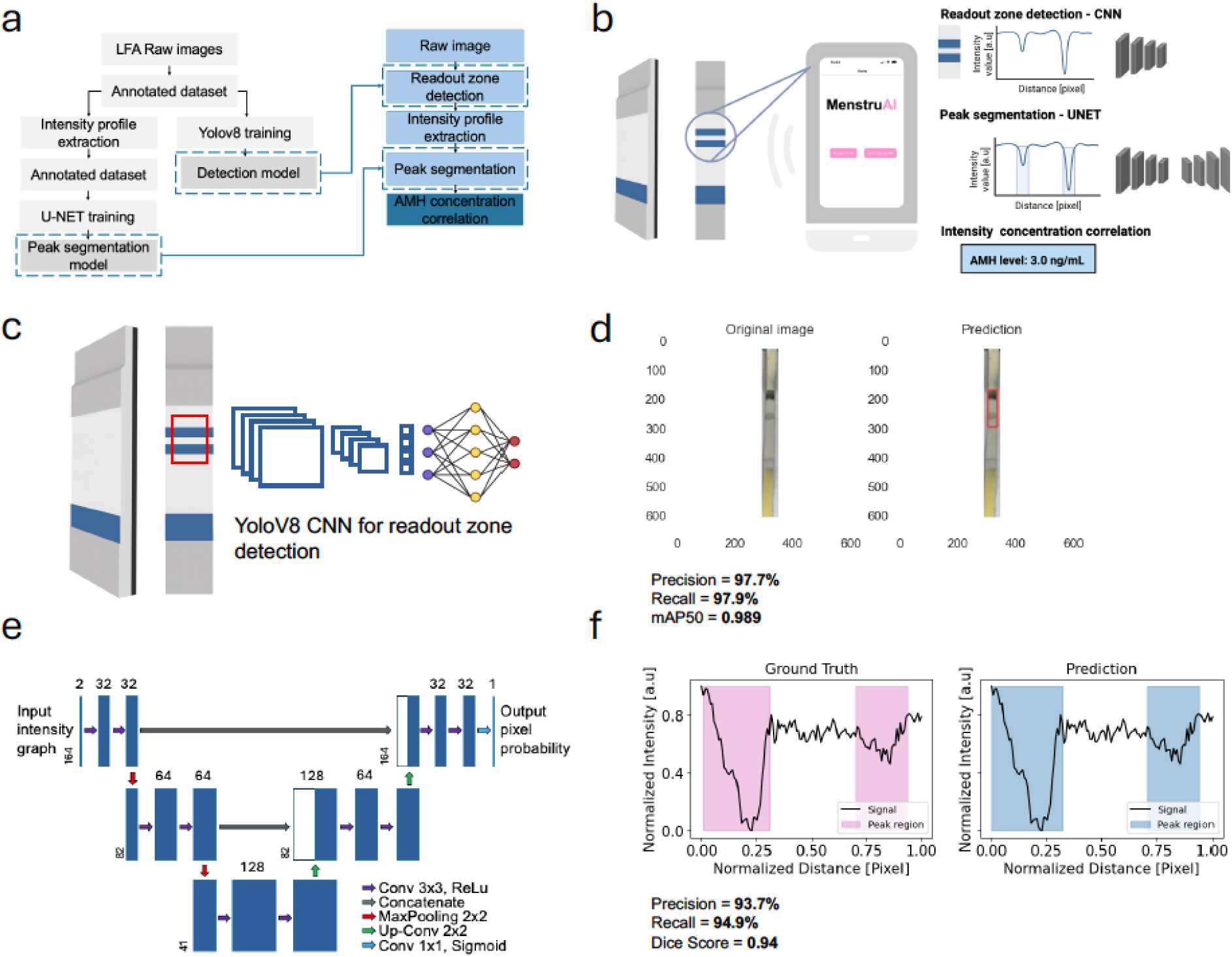
Machine learning based image analysis for AMH detection on lateral flow assay. a) Workflow of machine learning model dataset preparation and training. For the training process, a dataset was first generated using a set of raw images of LFA strips, manually labeled. This dataset was then used for the training of two ML models for readout zone detection and classification. Additionally, intensity profiles were extracted from the same images and manually annotated for peak and background regions. This dataset was then used for the training of the encoder decoder model to perform automatic peak segmentation on new (unseen) data. b) Concept of the smartphone-based machine learning (ML) for LFA test analysis. A test image was captured with a smartphone. An ML model was employed to identify the readout zone and classify the result, analyzing the pixel intensity profile to locate peaks, corresponding to control and test line intensities. These intensities can be directly linked to biomarker concentration, based on pre-established integrated calibration. c) Pre-trained convolutional neural network YOLOV8 used for automatic readout zone detection from a smartphone image of a LFA strip. d) Readout zone detection using YOLOV8. The CNN model automatically detected the region of interest where the readout of the test was located. The model reached a precision of 97.7%, Recall of 97.9% and mAP50 of 0.989 after training. e) Encoder-decoder model following a 1D U-NET architecture used for automatic peak segmentation of a pixel-intensity graph to extract the value of the test-line peak. f) Examples of peak segmentation on intensity profiles. The encoder-decoder model accurately detected the regions corresponding to a peak of intensity on the LFA strips. The model reached a precision of 93.7%, Recall of 94.9% and a dice score of 0.94.

### Detection and semi-quantification of anti-müllerian hormone in menstruation blood

To evaluate the capacity of our sensor to accurately detect and track AMH in menstruation blood, menstruation blood samples were obtained from healthy volunteers and used unprocessed. For diagnostic benchmarking, the samples were obtained and tested on the same day using a chemiluminescent assay at standard clinical chemistry laboratory. The menstrual blood samples were tested at the same time using the optimized AMH LFA. Additionally, feasibility and wearability were evaluated at the same time by asking the volunteers to wear a modified hygiene pad with the embedded LFA sensor and to use a dipstick-assay format on the blood collected from a menstrual cup. In all cases, the collected menstrual blood was used directly on the LFA without any preprocessing, and images of the LFA result were analyzed with a manual pixel-intensity analysis and with the machine-learning based analysis pipeline described above. The results obtained with the two methods were compared with the results obtained by the clinical chemistry laboratory (Figure 4a).

**Figure 4:**
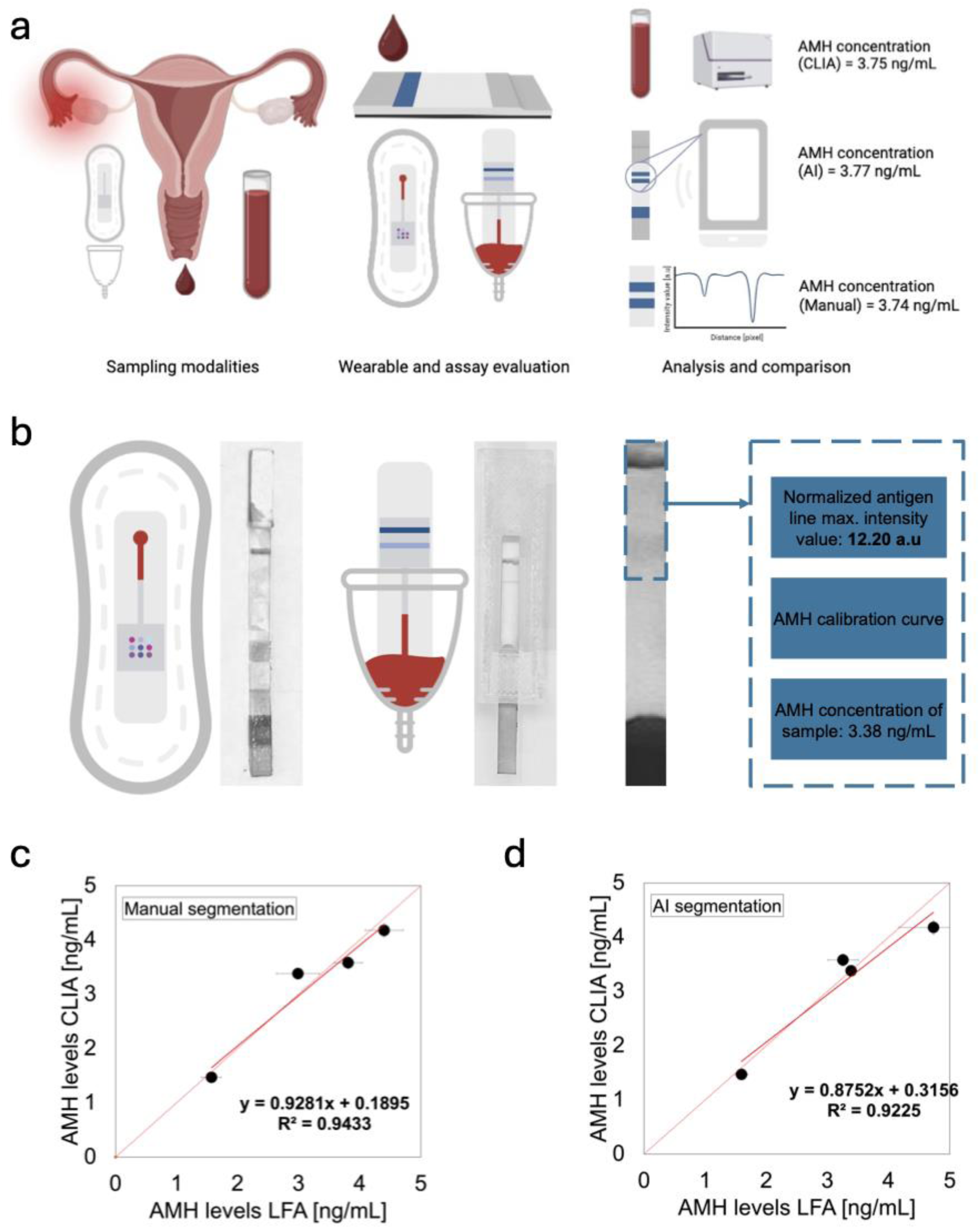
Detection of anti-müllerian hormone in unknown samples of menstruation blood (N=4) and comparison with CLIA methods. a) Sampling and testing protocols for the detection of AMH in menstruation blood samples. Menstruation blood samples were collected and analyzed using the gold-standard clinical chemistry testing. The samples were handed over for testing using the developed AMH LFA on the same day. Images of the test result were obtained using a smartphone. Manual segmentation using ImageJ was performed, along with detection and quantification of AMH in the sample using the AI-based image analysis software. b) Examples of AMH LFA integration strategies into sanitary pad-based wearable or from menstrual cup sampling with resulting test strips after user testing and example of quantification of AMH in a menstrual blood sample using the AI-based image analysis (images in grayscale). c) Performance of the AMH LFA versus CLIA methods using manual image analysis. d) Performance of the AMH LFA versus CLIA methods using AI-based image analysis.

The successful line development in the two sampling modalities confirmed that the LFA platform was fully functional when integrated into different end-use formats, facilitating reliable testing when users are given clear collection guidelines to preserve the sample in physiological conditions. In all cases, the generated LFA strips could be reliably analyzed (Figure 4b). Using manual image analysis by a trained user, the measured AMH values showed strong agreement with the expected concentrations (Figure 4c), with coefficient of variation (CV) values ranging from 6.03% to 11.70% (Table S1).

Notably, the mean deviation from expected values was within acceptable clinical limits (*40–42*) for point-of-care devices, supporting the reliability of the LFA system. We further assessed the performance of our AI-based image analysis pipeline. The AI-predicted AMH concentrations remained consistent with those from manual analysis (Figure 4d), with CVs between 0.63% and 11.95%. For instance, Sample 3, with an expected concentration of 3.38 ng/mL, was precisely measured by the AI (3.38 ng/mL, CV 0.63%) (Table 1), highlighting the robustness and reproducibility of the automated approach. The AI method also achieved comparable or better precision than manual analysis in certain cases (e.g., Sample 3 and Sample 5), demonstrating that our algorithm can match the performance of trained human users. Overall, these findings confirm that our LFA provides results in close concordance with standard clinical chemistry assays. The low and consistent CVs across both evaluation methods reinforce the robustness of our detection and analysis pipeline. These results support the feasibility of deploying a fully automated, smartphone-compatible platform for home-based ovarian health monitoring. It underscores the potential of our AI-integrated LFA to provide reliable, semi-quantitative AMH assessment from unprocessed menstrual blood.

Taken together, these results demonstrate that the AMH LFA delivers accurate and reproducible semi-quantitative measurements directly from fresh menstrual blood collected under realistic user conditions. The LFA sensor is versatile and can be adapted to different end-use formats and can therefore offer an easy, reliable and scalable alternative for point-of-care quantification of AMH.

## CONCLUSIONS

To conclude, we introduce a lateral flow assay (LFA) capable of detecting AMH directly in unprocessed menstrual blood, marking a significant step toward non-invasive, at-home ovarian health monitoring. By combining covalently conjugated 150 nm gold nanoshells with optimized assay chemistry and a machine learning-assisted image analysis pipeline integrated into smartphone app for ease-of-use, we achieved accurate and reproducible AMH quantification within the clinically relevant range in human (menstruation) blood. The platform demonstrated strong concordance with standard CLIA assays, with coefficients of variation that support its diagnostic potential. Notably, our AI-based interpretation method matched or exceeded manual analysis by trained users, reducing inter-operator variability and enabling reliable results from smartphone images. We demonstrated the feasibility and clinical validity of detecting AMH in unprocessed menstrual blood using a robust paper-based lateral flow platform. Beyond its application in menstruation blood, the platform can be readily adapted to finger prick blood testing, expanding its relevance also to male population for which AMH is an important biomarker of Sertoli cell function and certain gonadal disorders, highlighting that the presented framework holds diagnostic value beyond female fertility alone.(*43–45*) The assay’s flexible design makes it suitable for implenentation into both wearable (sanitary pad) and non-wearable (dipstick) formats, depending on the intended use case and user preference. This work highlights the transformative potential of (menstrual) blood-based AMH analysis as a scalable, user-friendly tool for empowering women in monitoring and managing their reproductive health. Future work will aim at expanding clinical validation, assess real-world usability across broader populations and investigate longitudinal monitoring.

## MATERIAL AND METHODS

### Materials

Sodium citrate dihydrate HOC(COONa)(CH_2_COONa)_2_·2H_2_O, HAuCl_4_, Phosphate-buffered saline, bovine serum albumin, sucrose C_12_H_22_O_11_, tween-20, NaH_2_PO_4_·H_2_O, Na_2_HPO_4_, Na_2_B_4_O_7_·10H_2_O, H_3_BO_3_, were purchased from Sigma-Aldrich. Anti-AMH detection and capture monoclonal antibodies (anti-AMH abx019327 and anti-AMH abx019316) were purchased from Lubioscience. Goat anti-mouse IgG were purchased from Lubioscience. AMH (abx060302) was purchased from Lubioscience. Silica-core 150 nm gold nanoshells were purchased from Nanocomposix. 1-ethyl-3-(3-dimethylaminopropyl) carbodiimide (EDC), N-Hydroxysuccinimide (NHS), and potassium phosphate reagent were obtained as part of a covalent conjugation kit for Silica-core 150 nm gold nanoshells (NanoComposix #GSZR150) obtained from Nanocomposix. Human serum was purchased from Sigma-Aldrich. All buffers and reagent solutions were prepared using distilled water or MiliQ water (>18.2Ø.cm, Milipore). The sample, conjugate, and absorbent pads, as well as nitrocellulose membranes and adhesive backing cards were obtained as part of a LFA assembly kit purchased from Nanocomposix. Additionally, Fusion 5 blood separation membrane was purchased from Cytiva.

### Human venous and menstruation blood samples

Before testing unprocessed menstruation blood, human serum (Sigma) was initially used for the assays. For each assay, spiked serum samples were tested to establish calibration curves for semi-quantitative biomarker detection. After demonstrating feasibility with serum samples, the assay was used with unprocessed human whole venous blood obtained from different volunteers for recovery experiments, to validate the calibration curves previously obtained (Table S2), before testing menstruation blood samples from volunteers and comparing with standard clinical laboratory testing (Chemiluminescent immunoassay using UniCel Dxl 800 Beckman Coulter). In total, ten healthy volunteers were enrolled for menstruation blood sampling on day 1 and 2 of menses and to evaluate the device usability and workflow. A subset of samples was used for the quantitative comparison. One of the samples (sample #4) was removed from the analysis due to hemolysis as confirmed by the clinical chemistry laboratory. Menstruation blood was collected by the volunteers using a menstrual cup (approved by ethics commission of the canton of St. Gallen, Switzerland, BASEC No. 2023-00562) and was donated to a gynecologist at the Cantonal Hospital St.

Gallen. The samples were tested on the same day using a chemiluminescent assay at standard clinical chemistry laboratory and using the optimized AMH LFA. Additionally, volunteers were asked to wear the modified hygiene pad with the embedded LFA sensor in the same conditions as they would with a commercially available sanitary pad, with no other restrictions or to use the dipstick-assay format on blood from the menstrual cup before sending a photo of the completed LFA for analysis. All samples, data and images of the completed tests were handed over to the research lab in an anonymous way (not transferring donor information). The collection of venous (BASEC No. 2016-00816) and menstruation blood (BASEC No. 2023-00562) was approved by the cantonal ethics commission of the Canton of St. Gallen, Switzerland.

### Reagents preparation

Phosphate buffer (0.01M, pH 7.4) was made using Na_2_HPO_4_ and NaH_2_PO_4_.H_2_O in MiliQ water. Borate buffer (100mM, pH9) was made using Na_2_B_4_O_7_·10H_2_O and H_3_BO_3_ in MiliQ water. The sample pad buffer consisted of a solution of PBS containing 0.5% (w/v) BSA and 1% (v/v) tween-20. Finally, the conjugate pad buffer in which nanoparticles were resuspended after conjugation consisted of a solution of PBS containing 5% (w/v) sucrose, 1% (w/v) BSA and 0.5% (v/v) tween-20.

### Apparatus

A Hidex sense 425-301 plate reader was used for the absorbance measurement and verification of nanoparticles conjugation. An iPhone 13 along with ImageJ software analysis were used for quantitative evaluation of the assay’s results.

### Gold nanoparticle synthesis

To synthesize gold nanospheres, a 60 mL solution of 2.2 mM of sodium citrate dihydrate was prepared in 3 Neck round bottom glass flask and brought to boiling point. 400 µL of a 25 mM HAuCl_4_ was added afterwards, resulting in seed nanoparticles of approximately 6 to 10 nm diameter after 15 min. Afterwards, a growth step was realized adding 400 µL of a 60 mM sodium citrate dihydrate solution the mix, followed by 400 µL of a 25 mM HAuCl_4_ solution. After cooling down, gold nanoparticles of approximately 20 nm diameter were fully synthesized.

### Conjugation of antibodies to gold nanoparticles

To prepare gold nanoparticle conjugates, the desired detection antibodies were passively conjugated to the surface of the 20 nm gold nanoparticles and 150 nm gold nanoshells from non-covalent interaction. Briefly, a solution of 20 nm AuNP at pH9 was mixed with a 100 µg/mL solution of anti-AMH antibody diluted in MiliQ water, followed by the addition of 0.25% BSA in MiliQ water for the obtention of Anti-AMH 20 nm AuNP conjugates. A solution of 150 nm AuNS at pH9 was mixed with a 400 µg/mL solution of anti-AMH antibody diluted in MiliQ water, followed by the addition of 0.1% BSA in MiliQ water for the obtention of Anti-AMH 150 nm AuNS conjugates. After incubation, the 20 nm gold nanoparticle solution was centrifuged at 10,000 rpm, and the 150 nm gold nanoshells solution was centrifuged at 2,000 rcf. Both were resuspended in the conjugate pad buffer. To prepare the 150 nm AuNS covalently conjugated with anti-AMH antibodies,

The covalent conjugation kit - BioReady Gold Nanoshells (AuNS) was used. EDC and sulfo-NHS were used to activate the surface of the carboxyl-functionalized AuNS by forming amide bonds that link carboxyl groups to primary amines on the antibody. Different concentrations of anti-AMH detection antibody were then incubated for 1h, 2h and overnight. After incubation, the 150 nm AuNS solution was washed two times with phosphate buffer, and finally it was centrifuged at 2000 rcf for 5 min and resuspended in the conjugate pad buffer.

### Lateral flow assay preparation

The LFA sensor consists of four components: a sample pad (blood filtration pad or not), a nitrocellulose membrane, a conjugate pad, and an absorbent pad. The sample pad was pre-treated with a sample pad buffer to assist in the fluid migration. The desired conjugated nanoparticles in a conjugate pad buffer were deposited on the conjugate pad. The absorbent pad was left untreated. A line of a 1 mg/mL anti-AMH capture antibody solution in phosphate buffer was deposited on the NC membrane to act as the test-line, and a 1.0 mg/mL goat anti-mouse IgG solution in phosphate buffer was deposited to act as the control line. All components were dried for 1h in a 37°C drying oven and later assembled on a plastic adhesive backing card. Each segment overlapped its neighboring one by 2 mm. Finally, individual LFA were cut to obtain regular paper-based sensors of 5 mm width and 6 cm length. The LFA strips were cut manually using a guillotine cutter (Paintersisters Lever cutter A4 compact, purchased from Galaxus.ch).

### Lateral flow optimization

The selection of antibody concentration, pH of nanoparticle solutions, and nanoparticle concentrations were optimized for the AMH LFA with 20 nm AuNP and 150 nm AuNS passively conjugated, using a nanoparticle-bioreceptor aggregation test to determine the optimal pH and minimum antibody concentration for complete nanoparticle coverage (Figure S1). Nanoparticle concentration was optimized by varying the optical density (OD) of AuNP solutions applied to the conjugate pad. LFA strips were prepared in triplicate, and the optimal signal intensity and sensitivity determined the final concentration. For the AMH LFA using 150 nm AuNS covalently conjugated, the assay was optimized following the recommendation of Nanocomposix. Different antibody concentrations, as well as different incubation times were tested. The choice was made after comparing signal intensity and sensitivity to ensure maximum assay performance. Finally, the comparison between AMH LFA using 20 nm AuNP and AMH LFA using 150 nm AuNS was based on signal intensity, sensitivity, and signal to noise ratio. The limit of detection for the detection of AMH was extrapolated from the linear regression using the limit of the blank (𝜇_𝑏𝑙𝑎𝑛𝑘_ + 3 ∗ 𝜎_𝑏𝑙𝑎𝑛𝑘_). Signal-to-noise ratio (SNR) were calculated using the mean intensity and the standard deviation of the noise.

### Test procedure

Once LFA strips were prepared, serum samples were spiked with AMH at desired concentrations to establish the calibration curves, whole blood samples were spiked with AMH at desired concentrations for the recovery experiments. Finally, menstruation blood samples were used unprocessed. 75 µL of sample were deposited on the sample pad before fully migrating towards the absorbent pad. The volume corresponds to the adequate amount of fluid necessary to initiate the LFA by capillary action while preventing overflow of the paper-based test. Images of the completed LFA were analyzed with ImageJ. The intensity profile of individual color channels of the LFA strip image was obtained, and the peak intensity of the lines was recorded. The intensity value was obtained by subtracting the peak value of the line from the background value. All measurements were triplicated. Images of the completed LFA obtained from volunteers and using unprocessed menstrual blood samples were analyzed in two ways: i) using manual image analysis of the pixel intensity profile with ImageJ to extract the value of the test line intensity peak, and obtain the AMH concentration with the pre-determined calibration curve, and ii) using an AI-based software method which automatically detected the readout zone from a smartphone image, processed the pixel intensity profile to segment the intensity peak, and provided the value of AMH concentration in the samples.

### Artificial intelligence assisted analysis using smartphone app

The smartphone-based analysis using AI has been built similarly as described in our previous work (*37*). Two distinct machine learning models were used to i) detect the readout zone and ii) perform peak segmentation on the intensity profile of the LFA strip to correlate the test line intensity to the biomarker concentration. The readout zone on the LFA is automatically detected first, and the pixel intensity profile of the appropriate region is obtained. The value of the test line intensity peak is extracted and is later used to correlate with the concentration of AMH in the tested sample using the calibration curves.

For the first model, a dataset containing 482 images of individual strips of AMH LFA was built. The dataset contained images taken under different lighting conditions and angles, and data augmentation was applied. The dataset has been manually labelled to draw a bounding box containing the AuNS-based colorful lines on each individual strip. The dataset was divided into training, testing and validation sets representing 70%, 20% and 10% respectively. Transfer learning has been leveraged, using a pre-trained yoloV8 convolutional neural network(*46*) model to learn the detection and segmentation of the readout zone using 10 epochs and a batch size of 10.

For the second model, the automatic peak segmentation was performed using an encoder-decoder model following a 1D U-NET architecture(*47*). To generate the training dataset, the intensity profiles from the 789 LFA strips were manually extracted. They were manually labelled, attributing a probability 1 to the pixels displaying a colorful line on the LFA and a probability 0 to the pixels consisting of the background region on the LFA. The final dataset was divided into training and testing sets representing 80% and 20% and the encoder-decoder model was trained using the pixel position and the corresponding intensity as input for supervised learning of the probability of the pixel to fall within a peak region or a background region. The model was built with a contracting encoder path consisting of 3 blocks of convolutional layers, and an expanding decoder path consisting of 2 blocks of up sampling and convolutional layers. The sigmoid activation function was used for the output layer. Skip connections were used to link the encoder and decoder blocks at each level. The model was trained using a bespoke loss function combining dice loss for class imbalance and segmentation overlap and binary cross entropy loss for pixel-wise binary classification. The model was trained over 200 epochs and batch size of 8.

## Data Availability

The data that support the findings of this study are available from the corresponding author upon reasonable request.

## ACKNOWLEDGEMENTS

We thank the anonymous menstruation blood donors for their participation in this study, and Oscar Cipolato for proofreading and providing feedback on the manuscript. We acknowledge funding by ETH Zurich and in parts by the Swiss National Science Foundation (Eccellenza grant no. 181290, I.K.H) and the Vontobel Foundation.

## CONFLICT OF INTEREST

Lucas Dosnon and Inge K. Herrmann declare inventorship on a patent application filed by ETH Zurich and Empa on the in-pad menstruation blood analysis platform (L. Dosnon, I.K. Herrmann, A device for detection of biomarkers, EP23209241). The authors declare no other competing interests.

## AUTHOR CONTRIBUTIONS

Conceptualization: L.D, T.R, I.K.H, Methodology: L.D, T.R, I.K.H, Investigation: L.D, S.S.A, Visualization: L.D, S.S.A Funding acquisition: I.K.H, Project administration: L.D, I.K.H, Supervision: I.K.H, Writing—original draft: L.D, I.K.H, Resources: T.R, I.K.H, Validation: L.D, Writing—review and editing: L.D, I.K.H, Data curation: L.D, Formal analysis: L.D, Software: L.D.

## Notes

### Author Declarations

The collection of venous (BASEC No. 2016-00816) and menstruation blood (BASEC No. 2023-00562) was approved by the cantonal ethics commission of the Canton of St. Gallen, Switzerland.

## REFERENCES

1. O. Oktem, K. Oktay, The Ovary. Ann. N. Y. Acad. Sci. 1127, 1–9 (2008).

2. G. F. Erickson, NORMAL OVARIAN FUNCTION. Clin. Obstet. Gynecol. 21, 31 (1978).

3. H. Peters, K. P. McNatty, The Ovary: A Correlation of Structure and Function in Mammals (University of California Press, 1980).

4. G. C. Jayson, E. C. Kohn, H. C. Kitchener, J. A. Ledermann, Ovarian cancer. The Lancet 384, 1376–1388 (2014).

5. U. A. Matulonis, A. K. Sood, L. Fallowfield, B. E. Howitt, J. Sehouli, B. Y. Karlan, Ovarian cancer. Nat. Rev. Dis. Primer 2, 16061 (2016).

6. R. J. Norman, D. Dewailly, R. S. Legro, T. E. Hickey, Polycystic ovary syndrome. The Lancet 370, 685–697 (2007).

7. S. J. Chon, Z. Umair, M.-S. Yoon, Premature Ovarian Insufficiency: Past, Present, and Future. Front. Cell Dev. Biol. 9 (2021).

8. P. Vercellini, P. Viganò, E. Somigliana, L. Fedele, Endometriosis: pathogenesis and treatment. Nat. Rev. Endocrinol. 10, 261–275 (2014).

9. R. Azziz, E. Carmina, Z. Chen, A. Dunaif, J. S. E. Laven, R. S. Legro, D. Lizneva, B. Natterson-Horowtiz, H. J. Teede, B. O. Yildiz, Polycystic ovary syndrome. Nat. Rev. Dis. Primer 2, 16057 (2016).

10. S. K. Agarwal, C. Chapron, L. C. Giudice, M. R. Laufer, N. Leyland, S. A. Missmer, S. S. Singh, H. S. Taylor, Clinical diagnosis of endometriosis: a call to action. Am. J. Obstet. Gynecol. 220, 354.e1–354.e12 (2019).

11. J. M. Liberto, S.-Y. Chen, I.-M. Shih, T.-H. Wang, T.-L. Wang, T. R. Pisanic, Current and Emerging Methods for Ovarian Cancer Screening and Diagnostics: A Comprehensive Review. Cancers 14, 2885 (2022).

12. M.-K. Hong, D.-C. Ding, Early Diagnosis of Ovarian Cancer: A Comprehensive Review of the Advances, Challenges, and Future Directions. Diagnostics 15, 406 (2025).

13. N. di Clemente, C. Racine, A. Pierre, J. Taieb, Anti-Müllerian Hormone in Female Reproduction. Endocr. Rev. 42, 753–782 (2021).

14. Anti-Mullerian hormone. 10.1152/physrev.1986.66.4.1038.

15. S. Tsepelidis, F. Devreker, I. Demeestere, A. Flahaut, Ch. Gervy, Y. Englert, Stable serum levels of anti-Müllerian hormone during the menstrual cycle: a prospective study in normo-ovulatory women. Hum. Reprod. 22, 1837–1840 (2007).

16. W. J. K. Hehenkamp, C. W. N. Looman, A. P. N. Themmen, F. H. de Jong, E. R. te Velde, F. J. M. Broekmans, Anti-Müllerian Hormone Levels in the Spontaneous Menstrual Cycle Do Not Show Substantial Fluctuation. J. Clin. Endocrinol. Metab. 91, 4057–4063 (2006).

17. L. M. E. Moolhuijsen, J. A. Visser, Anti-Müllerian Hormone and Ovarian Reserve: Update on Assessing Ovarian Function. J. Clin. Endocrinol. Metab. 105, 3361–3373 (2020).

18. C. Peluso, F. L. A. Fonseca, I. F. Rodart, V. Cavalcanti, G. Gastaldo, D. M. Christofolini, C. P. Barbosa, B. Bianco, AMH: An ovarian reserve biomarker in assisted reproduction. Clin. Chim. Acta 437, 175–182 (2014).

19. F. J. Broekmans, J. A. Visser, J. S. E. Laven, S. L. Broer, A. P. N. Themmen, B. C. Fauser, Anti-Müllerian hormone and ovarian dysfunction. Trends Endocrinol. Metab. 19, 340–347 (2008).

20. K. P. Tremellen, M. Kolo, A. Gilmore, D. N. Lekamge, Anti-müllerian hormone as a marker of ovarian reserve. Aust. N. Z. J. Obstet. Gynaecol. 45, 20–24 (2005).

21. Full article: New AMH assay allows rapid point of care measurements of ovarian reserve. https://www.tandfonline.com/doi/full/10.1080/09513590.2017.1306735#abstract.

22. R. Tal, D. B. Seifer, M. Khanimov, H. E. Malter, R. V. Grazi, B. Leader, Characterization of women with elevated antimüllerian hormone levels (AMH): correlation of AMH with polycystic ovarian syndrome phenotypes and assisted reproductive technology outcomes. Am. J. Obstet. Gynecol. 211, 59.e1–59.e8 (2014).

23. A. La Marca, A. Volpe, Anti-Müllerian hormone (AMH) in female reproduction: is measurement of circulating AMH a useful tool? Clin. Endocrinol. (Oxf.) 64, 603–610 (2006).

24. D. Dewailly, C. Y. Andersen, A. Balen, F. Broekmans, N. Dilaver, R. Fanchin, G. Griesinger, T. W. Kelsey, A. La Marca, C. Lambalk, H. Mason, S. M. Nelson, J. A. Visser, W. H. Wallace, R. A. Anderson, The physiology and clinical utility of anti-Mullerian hormone in women. Hum. Reprod. Update 20, 370–385 (2014).

25. D. Gassner, R. Jung, First fully automated immunoassay for anti-Müllerian hormone. Clin. Chem. Lab. Med. CCLM 52, 1143–1152 (2014).

26. G. Demirdjian, S. Bord, C. Lejeune, R. Masica, D. Rivière, L. Nicouleau, P. Denizot, P.-Y. Marquet, Performance characteristics of the Access AMH assay for the quantitative determination of anti-Müllerian hormone (AMH) levels on the Access* family of automated immunoassay systems. Clin. Biochem. 49, 1267–1273 (2016).

27. R. Punchoo, S. Bhoora, Variation in the Measurement of Anti-Müllerian Hormone – What Are the Laboratory Issues? Front. Endocrinol. 12, 719029 (2021).

28. Q. Cai, S. Jin, H. Zong, L. Pei, K. Cao, L. Qu, Z. Li, A Quadruplex Ultrasensitive Immunoassay for Simultaneous Assessment of Human Reproductive Hormone Proteins in Multiple Biofluid Samples. Anal. Chem. 95, 11641–11648 (2023).

29. Y. Li, B. Luo, Y. Liu, S. Wu, S. Shi, H. Chen, M. Zhao, Microfluidic immunosensor based on a graphene oxide functionalized double helix microfiber coupler for anti-Müllerian hormone detection. Biomed. Opt. Express 14, 1364–1377 (2023).

30. Y. Wang, E. E. Dzakah, Y. Kang, Y. Cai, P. Wu, Y. Cui, Y. Huang, X. He, Development of anti-Müllerian hormone immunoassay based on biolayer interferometry technology. Anal. Bioanal. Chem. 411, 5499–5507 (2019).

31. A. Nandi, S. W. Gillespie, S. N. Ice, D. Thakur, Y. K. Gaur, K. Singh, K. Sharma, Electrochemical immunosensors targeting hormonal biomarkers for polycystic ovary syndrome: Recent advances and analytical bottlenecks. Biosens. Bioelectron. X 26, 100657 (2025).

32. S. Naseri, K. Lerma, P. Blumenthal, Comparative Assessment of Serum versus Menstrual Blood for Diagnostic Purposes: A Pilot Study. *J*. Clin. Lab. Med. 4 (2019).

33. S. Naseri, Y. Rosenberg-Hasson, H. T. Maecker, M. I. Avrutsky, P. D. Blumenthal, A cross-sectional study comparing the inflammatory profile of menstrual effluent vs. peripheral blood. Health Sci. Rep. 6, e1038 (2023).

34. S. A. Cederholm-Williams, M. C. Rees, A. C. Turnbull, Examination of certain coagulation factors in menstrual fluid from women with normal blood loss. Thromb. Haemost. 52, 224–225 (1984).

35. S. Naseri, S. Young, G. Cruz, P. D. Blumenthal, Screening for High-Risk Human Papillomavirus Using Passive, Self-Collected Menstrual Blood. Obstet. Gynecol. 140, 470–476 (2022).

36. S. Naseri, M. I. Avrutsky, C. Capati, K. Desai, R. Alvero, P. D. Blumenthal, Concordance of hemoglobin A1c and reproductive hormone levels in menstrual and venous blood. FS Rep., doi: 10.1016/j.xfre.2023.11.009 (2023).

37. L. Dosnon, T. Rduch, C. Meyer, I. K. Herrmann, A Wearable In-Pad Diagnostic for the Detection of Disease Biomarkers in Menstruation Blood. Adv. Sci. n/a, e05170.

38. S. Lee, S. Kim, D. S. Yoon, J. S. Park, H. Woo, D. Lee, S.-Y. Cho, C. Park, Y. K. Yoo, K.-B. Lee, J. H. Lee, Sample-to-answer platform for the clinical evaluation of COVID-19 using a deep learning-assisted smartphone-based assay. Nat. Commun. 14, 2361 (2023).

39. N. C. K. Wong, S. Meshkinfamfard, V. Turbé, M. Whitaker, M. Moshe, A. Bardanzellu, T. Dai, E. Pignatelli, W. Barclay, A. Darzi, P. Elliott, H. Ward, R. J. Tanaka, G. S. Cooke, R. A. McKendry, C. J. Atchison, A. A. Bharath, Machine learning to support visual auditing of home-based lateral flow immunoassay self-test results for SARS-CoV-2 antibodies. Commun. Med. 2, 1–10 (2022).

40. A. Heydecke, K. Gullsby, Evaluation of the performance of a rapid antigen test (Roche) for COVID-19 diagnosis in an emergency setting in Sweden. J. Med. Virol. 95, e28537 (2023).

41. Assessing the Reliability of Commercially Available Point of Care in Various Clinical Fields. https://openpublichealthjournal.com/VOLUME/12/PAGE/342/FULLTEXT/.

42. E. K. Harris, Statistical principles underlying analytic goal-setting in clinical chemistry. Am. J. Clin. Pathol. 72, 374–382 (1979).

43. D. Dewailly, J. Laven, AMH as the primary marker for fertility. Eur. J. Endocrinol. 181, D45–D51 (2019).

44. H.-Y. Xu, H.-X. Zhang, Z. Xiao, J. Qiao, R. Li, Regulation of anti-Müllerian hormone (AMH) in males and the associations of serum AMH with the disorders of male fertility. Asian J. Androl. 21, 109 (2018).

45. E. Matuszczak, A. Hermanowicz, M. Komarowska, W. Debek, Serum AMH in Physiology and Pathology of Male Gonads. Int. J. Endocrinol. 2013, 128907 (2013).

46. G. Jocher, A. Chaurasia, J. Qiu, Ultralytics YOLO, version 8.0.0 (2023); https://github.com/ultralytics/ultralytics.

47. “U-Net: Convolutional Networks for Biomedical Image Segmentation” in Lecture Notes in Computer Science (Springer International Publishing, Cham, 2015; http://link.springer.com/10.1007/978-3-319-24574-4_28), pp. 234–241.

