## Supplementary Information for "At Home Detection of Ovarian Health Biomarker in Menstruation Blood"

+41 (0)58 765 7153

| **Sample** | **Expected value [ng/mL]** | **Measured value (Manual) [ng/mL]** | **Cv (Manual) [%]** | **Measured value (AI) [ng/mL]** | **Cv (AI)[%]** |
| --- | --- | --- | --- | --- | --- |
| **Sample 1** | 3.58 | 3.81 | 6.03 | 3.25 | 7.58 |
| **Sample 2** | 4.18 | 4.40 | 7.04 | 4.73 | 11.95 |
| **Sample 3** | 3.38 | 2.99 | 11.70 | 3.38 | 0.63 |
| **Sample 5** | 1.47 | 1.53 | 10.19 | 1.59 | 6.59 |

**Table S1:** Results of the detection and quantification of AMH in menstruation blood samples (N=4) using manual image analysis and AI-based image analysis. (Sample #4 removed from analysis due to confirmed hemolysis.)

| **Spiked AMH [ng/mL]** | **Expected value [ng/mL]** | **Measured value [ng/mL]** | **Recovery [%]** |
| --- | --- | --- | --- |
| 10 | 14.18 | 12.63 | 89.11 |
| 10 | 10.38 | 9.64 | 92.89 |
| 5 | 9.18 | 10.75 | 117.11 |
| 5 | 5.38 | 4.47 | 83.11 |
| 4 | 8.18 | 9.32 | 114.05 |
| 3 | 7.18 | 8.02 | 111.83 |
| 2 | 6.18 | 6.98 | 112.96 |
| 0 | 0.38 | 0.29 | 77.55 |

**Table S2:** Recovery experiment results using human venous whole blood spiked with different concentrations of AMH.

**Figure S1:** Optimization of 20 nm and 150 nm gold nanoparticles solutions pH and antibody concentrations during nanoparticle-aggregation test for optimal conjugation and maximization of assay performances. a) Conjugation output at various pH of 20 nm gold nanoparticles solution at the determined optimal concentration of AMH antibody solution. b) Conjugation output at various concentrations of AMH antibody solution for the determined optimal pH of 20 nm gold nanoparticle solution. c) Conjugation output at various pH of 150 nm gold nanoshells solution at the determined optimal concentration of AMH antibody solution. d) Conjugation output at various concentrations of AMH antibody solution for the determined optimal pH of 150 nm gold nanoshells solution.
